# Compliance to Iron Folic Acid Supplementation and its associated factors among post-partum mothers of Bharatpur Metropolitan

**DOI:** 10.1101/2024.07.18.24310631

**Authors:** Amshu Pokhrel, Bimala Bhatta, Anup Adhikari

**Author notes:** Corresponding author (AP).

## Abstract

**Background:** Anemia is a major global health issue, especially affecting young children, pregnant and postpartum women, and adolescent girls, with 1.92 billion affected in 2021, notably in sub-Saharan Africa and South Asia. Iron deficiency is a key cause, requiring interventions like iron and folic acid supplementation. Despite efforts in Nepal, anemia prevalence fluctuates, with compliance a concern. This study assesses iron and folic acid supplementation compliance among postpartum mothers in Bharatpur Metropolitan City.

**Method:** A community-based cross-sectional study was conducted among 303 postpartum mothers with babies aged 45days to 1 year in Bharatpur Metropolitan City. Data were collected through face-to-face interviews using a pre-tested questionnaire.

**Results:** The compliance rate for iron and folic acid supplementation among postpartum mothers was 48.2%. Among different sociodemographic variables, religion of participants was found to be significantly associated with compliance to iron folic acid supplementation (OR 5.367, 95% CI 1.173-16.636). Moreover, participants having more than four antenatal visits (OR 3.465, 95% CI 1.366-8.792), participants having good knowledge about anemia (OR 5.554, 95% CI 2.485-12.415), and participants having good knowledge about iron and folic acid supplementation (OR 2.442, 95% CI 1.064-5.60) were also significantly associated with higher compliance.

**Conclusion:** To improve IFAS adherence, healthcare providers should focus on health education on the postpartum importance of iron and folic acid supplementation, along with diligent follow-up using culturally tailored strategies. Enhanced compliance will lead to better health outcomes for mothers and their babies.

## Introduction

Pregnancy heralds a transformative journey in a woman’s life, marked not only by the physical changes she undergoes but also by the increased demand for essential nutrients to sustain her health and that of her developing fetus [1]. Among these vital nutrients, iron and folic acid stand out as indispensable contributors to maternal well-being and fetal development. Deficiencies in iron can result in maternal anemia, characterized by fatigue, weakness, and impaired immune function, and can pose significant risks to both maternal and fetal health[2].

Anemia is a global public health problem affecting both developing and developed countries. It is the result of wide variety of causes, the most significant contributor to the onset of anemia is iron deficiency[3]. Pregnant women are particularly at the increased risk of IFA deficiency due to increases nutrient requirements[4]. Worldwide anemia prevalence data suggest that the normal dietary intake of iron is insufficient to meet daily requirement for a significant proportion of pregnant woman[5].Maternal anemia is a pressing issue in countries like Nepal, affecting 30-50% of pregnant women. Globally, iron deficiency, the main cause of anemia, contributes to half of all cases. This problem is notably significant among women of reproductive age, including postnatal women, leading to adverse outcomes for both mother and child. With a global impact of 38% in pregnant women and 29% in non-pregnant women, anemia is a major concern [6,7]. According to NDHS 2022, the prevalence of anemia in pregnant women is 33% in Nepal.

The World Health Organization recommends daily oral IFA supplementation for pregnant and postnatal women to prevent maternal anemia and related complications. Understanding the factor’s influencing compliance is crucial for designing targeted interventions and assessing the effectiveness of existing strategies in reducing anemia rates in this specific population. Despite government efforts, anemia persists as a public health problem, and the unsatisfactory compliance rate with IFA supplementation underscores the need for further investigation into influencing factors with only 60% of Nepalese women receiving a 180-day supply of Iron Folic Acid during pregnancy in the year 2078/79, and 52.6% of postpartum women receiving a 45-day supply according to annual health report of Department of Health Service of Fiscal Year 2077/78.

This research initiative seeks to address a crucial gap in existing literature by extending the focus beyond pregnancy and encompassing the often-overlooked postpartum phase. By investigating factors influencing IFA compliance, the study aims to contribute detailed insights that can guide customized interventions for both healthcare professionals and policymakers. The ultimate goal is to foster a more comprehensive and effective approach to IFA supplementation, promoting the well-being of mothers and children in Bharatpur Metropolitan City.

## Materials and Methods

### Study Design and Setting

A community-based cross-sectional study was carried out among postpartum mothers in Bharatpur Metropolitan City. Bharatpur Metropolitan City consist total of 29 ward and the study focused on randomly selected 8 wards which includes ward 4, 7, 11, 14, 15, 17, 18, and 26. Bharatpur, the only metropolitan area in the Chitwan District, encompasses both rural and urban populations. This unique blend offers a genuine representation of various demographics and lifestyles, making it an excellent setting for thorough research. The data was collected between April 28 and May 30 2024.

### Study Population and Sampling Procedure

Post-partum mothers having baby from 45 days to 1 year were included in the study and post-partum mothers having baby from 45 days to 1 year who were seriously ill and unable to communicate were excluded from the study.

The sample size was determined using Cochran’s formula for finite populations, considering a preceding prevalence of 69%. The margin of error was set at 0.05%, and the confidence interval was 95%. The finite population for the study was 3,853, obtained from HMIS data for the year 2078/79. Based on this, the sample size was calculated, and a total of 303 mothers were selected from 8 wards. These 8 wards were randomly chosen from a total of 29 wards. From each selected ward, a list of villages was compiled, and villages were selected randomly. A list of postpartum mothers with children aged 6 weeks to 1 year from each selected village was then obtained. Study participants were chosen from these randomly selected villages. Female Community Health Volunteers (FCHVs) played a key role in locating the respondents within the community.

### Study Variables

The study includes eight sociodemographic variables: age, religion, ethnicity, education, occupation, husband’s education, husband’s occupation, and monthly income. Obstetric health-related variables include: gravida, gestational age at first ANC, frequency of ANC visits, history of anemia, abortion, miscarriage, and pregnancy complications. It also considers knowledge of anemia and iron-folic acid (IFA) supplementation. Health facility-related variables include distance to the nearest facility, time to collect IFA, counseling received, and tablets provided per visit. Supplement-related variables include the number of tablets consumed, side effects, management of side effects, and time of day for consumption.

Compliance with Iron Folic Acid (IFA) is measured using two main questions: the consumption of IFA tablets during pregnancy and the consumption of IFA tablets during the post-partum period. For pregnancy, the consumption is recorded as the total number of tablets taken before delivery, with one tablet consumed daily starting from the fourth month until delivery. For the post-partum period, the consumption is recorded as the total number of tablets taken after delivery and up to 45 days, with one tablet consumed daily.

### Operational Definition

The dependent variable, compliance to iron folic acid supplementation refers to consistent consumption of 225 tablets, taken one per day starting from the second trimester of pregnancy, comprises 180 tablets during the antenatal period and 45 tablets postpartum, as prescribed according to government recommendations.

Post-partum mothers refers to the mothers having baby up to 1 year starting from 45 days The period from the child birth up to the one year of delivery is known as extended postpartum period [8].

Women’s knowledge of anemia was assessed by five multiple-choice questions asking about signs and symptoms of anemia, possible causes of anemia, groups susceptible to anemia, and prevention of anemia. The correct answer for each item was scored as "1," and the incorrect answer was scored as "0." The items were summed, and the mean was calculated. If the pregnant women score above the mean, they had good knowledge, and if the pregnant women score below the mean, they had poor knowledge regarding anemia [5].

Women’s knowledge of IFA supplementation was assessed by five questions asking about the benefits of IFA, frequency of use, duration of use, and side effects. The correct answer for each item was scored as "1," and the incorrect answer was scored as "0." The items were summed, and the mean was calculated. If the pregnant women score above the mean, they had good knowledge, and if the pregnant women score below the mean, they had poor knowledge regarding IFA supplementation [5].

Side effects of Iron Folic Acid (IFA) supplementation refer to any adverse and unwanted reactions experienced by individuals who have consumed IFA tablets, including symptoms such as vomiting, nausea, dizziness, heartburn, and diarrhea[9].

Distance to nearest health facility refers to the amount of time it takes for an individual to travel from their location to nearest health facility.

The average time to collect Iron Folic Acid (IFA) from a health facility refers to the mean duration it takes for individuals (typically pregnant women) to obtain their IFA supplements from a designated health facility.

Consumption time of day refers to the specific time during the day when an individual takes Iron Folic Acid tablets.

### Tools and Techniques of Data Collection

Data was collected through a face-to-face interviewer-administered questionnaire. The data was collected in printed questionnaire. The consistency and completeness of the data were checked after the completion of the data collection.

The questionnaires in this study were based on a thorough literature review. The tool was initially developed in English and later translated into the local Nepali language. For content validity, we consulted experts having similar experience. The tool was pre-tested among 31 participants in the neighboring ward and later adjusted and modified before the data collection.

### Data Processing and Analysis

The data was collected using printed questionnaires. The collected data was kept safely, and it was entered into epidata as soon as possible in order to maintain the safety of the data. The data were expert from the epidata to IBM SPSS Statistics 22 for further processing, including cleaning, coding, and analysis. Frequency, mean, and standard deviation were used for descriptive statistics. The Chi-square test, Bi-variate and multiple logistic regression analysis were performed. Variables that were significant in the chi-square test were included in the multicollinearity test. A multicollinearity test was performed using variance inflation factor (VIF), and variables less than 2 were included in the multivariate study. Furthermore, the goodness of fit for the final logistic model was tested using the Hosmer and Lemeshow test.

### Ethical Considerations

The Ethical approval from the Institutional Review Committee (IRC) of Pokhara University was obtained (reference number 141/2080/81). Written approval to conduct the study was also acquired from Bharatpur Metropolitan City. Written informed consent was taken from each of the participants before the interview and confidentiality of the participants was maintained.

## Results

### Sociodemographic characteristics of Participants

Table 1 presents the sociodemographic characteristics of the participants. Over two-thirds (67%) of participants were aged 25-29, with a mean age of 28.83 and standard deviation of 2.84years. Half were Brahmin (50%), 31% were Janjati, and 82% were Hindu. Nearly half (45%) had completed secondary education, and 58% were homemakers. Among husbands, 36% had secondary education, and 25% worked in agriculture. Additionally, 42% of households had an income below Rs.10,000, with a similar proportion earning Rs.30,000.

**Table 1:** Sociodemographic Characteristics of the respondents.

| Characteristics | Frequency | Percentage (%) |
| --- | --- | --- |
| **Age of the respondents** |  |  |
| **Mean age (Standard Deviation) :28.83 (2.84)** |  |  |
| 20-24 | 56 | 18.5 |
| 25-29 | 203 | 67.0 |
| 30-34 | 38 | 12.5 |
| 35-39 | 6 | 2.0 |
| **Ethnicity** |  |  |
| Dalit | 44 | 14.5 |
| Janajati | 96 | 31.7 |
| Madhesi | 2 | .7 |
| Muslim | 8 | 2.6 |
| Brahmin/Chettri | 153 | 50.5 |
| **Religion** |  |  |
| Hindu | 248 | 81.8 |
| Buddhist | 38 | 12.5 |
| Christ | 8 | 2.6 |
| Muslim | 9 | 3.0 |
| **Education** |  |  |
| Cannot read and write | 9 | 3.0 |
| Non formal education | 56 | 18.5 |
| Primary | 68 | 22.4 |
| Secondary | 135 | 44.6 |
| Bachelor | 31 | 10.2 |
| Masters | 4 | 1.3 |
| **Occupation** |  |  |
| Government employee | 8 | 2.6 |
| Private employee | 15 | 5.0 |
| Foreign employee | 2 | .7 |
| Homemaker | 176 | 58.1 |
| Agriculture | 60 | 19.8 |
| Student | 21 | 6.9 |
| Business | 18 | 5.9 |
| Unemployed | 3 | 1.0 |
| **Husband Education** |  |  |
| Cannot read and write | 5 | 1.7 |
| Non formal education | 54 | 17.8 |
| Primary | 75 | 24.8 |
| Secondary | 110 | 36.3 |
| Bachelor | 53 | 17.5 |
| Masters | 6 | 2.0 |
| **Husband Occupation** |  |  |
| Government employee | 35 | 11.6 |
| Private employee | 63 | 20.8 |
| Foreign employee | 43 | 14.2 |
| Agriculture | 80 | 26.4 |
| Daily labor | 23 | 7.6 |
| Student | 1 | .3 |
| Business | 52 | 17.2 |
| Unemployed | 6 | 2.0 |
| **Monthly Income** |  |  |
| **Median Income (IQR): 30000 (40000)** |  |  |
| ≤10000 | 130 | 42.9 |
| 10000-30000 | 44 | 14.5 |
| ≥30000 | 129 | 42.6 |

#### Obstetric Health related Characteristics of Participants

Table 2 shows the obstetric health related characteristics of participants. All women had their ANC visits, two fifth of the participants attending more than four times (40.3%). The first ANC visit is predominantly in the first month (44.2%), often occurring in health posts (40.9%) or government hospitals (28.1%). Blood tests during pregnancy are prevalent (89.1%), primarily conducted during the first visit (97.4%). Anemia history is reported by a small percentage (2.6%), managed primarily through consultation with doctors and iron tablet consumption. Miscarriages are reported by a minority (5.0%), while abortions are rare (0.7%). Low birth weight (LBW) baby history is minimal (1.0%). Complications during pregnancy are reported by 16.5%, with gestational diabetes being the most prevalent (36%). Postnatal care (PNC) is received by two fifth of the participants (40.6%), with half of them attending three times (51.2%). The majority report planned pregnancies (89.8%).

**Table 2:**
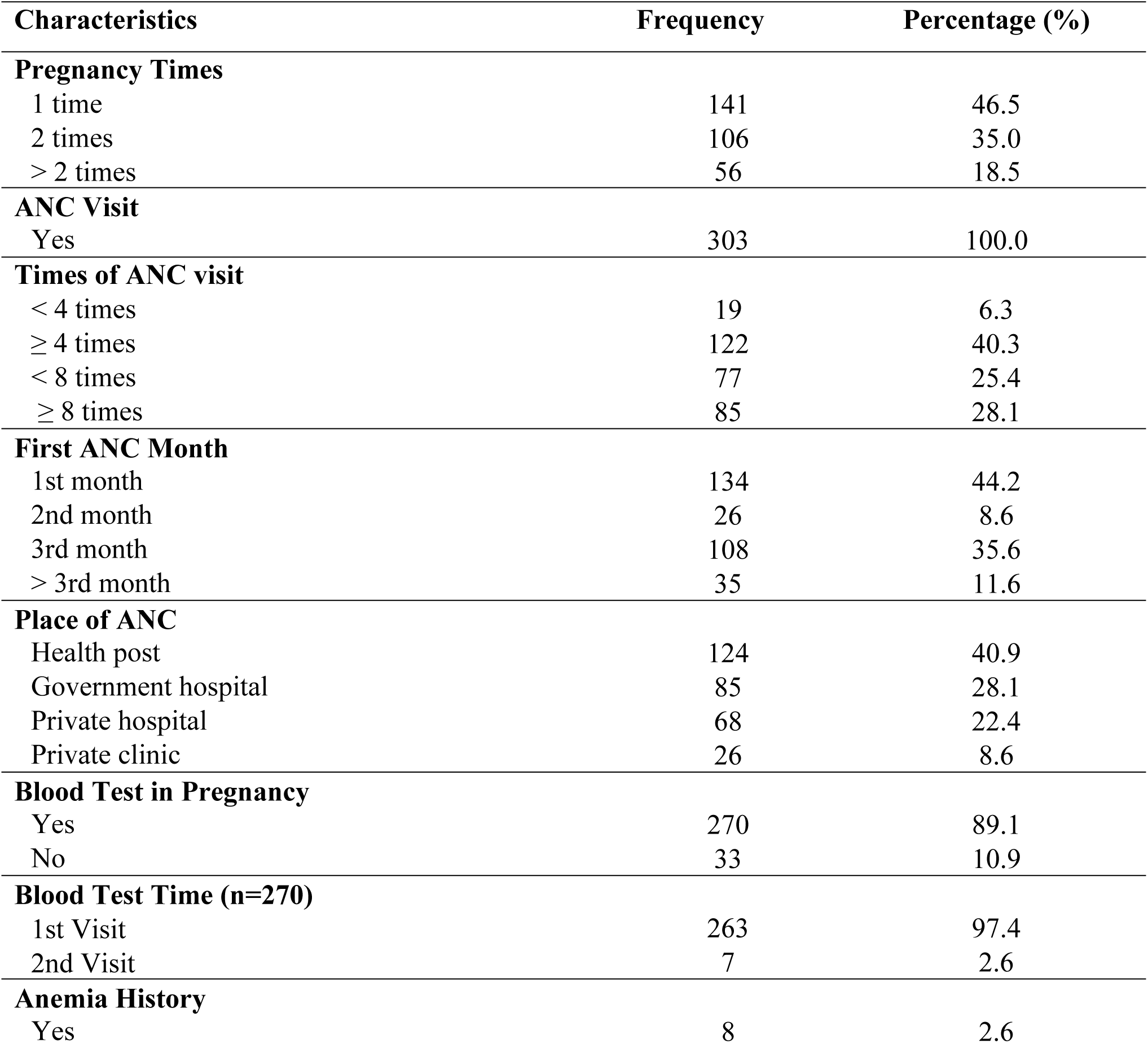
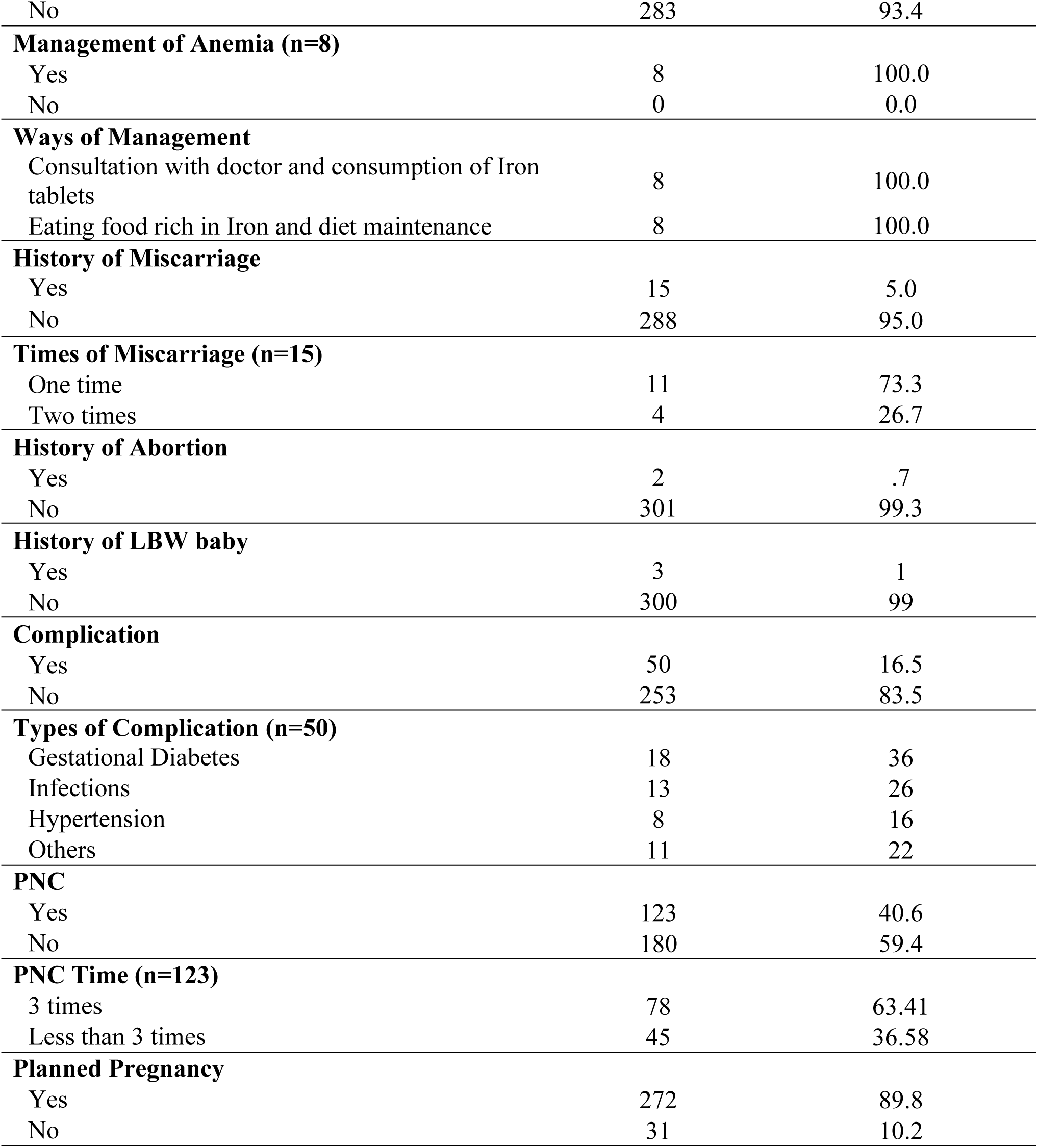
Obstetric health related characteristics of respondents.

### Knowledge on Anemia and IFA

Table 3 shows the knowledge of respondents regarding anemia and iron folic acid. More than half of the respondents had good knowledge on Anemia knowledge (56.1%) whereas regarding the knowledge of Iron Folic Acid two third of the respondents had good knowledge (60.4%).

| Characteristics | Frequency | Percentage (%) |
| --- | --- | --- |
| **Knowledge on Anemia** |  |  |
| Good Knowledge | 170 | 56.1 |
| Poor Knowledge | 133 | 43.9 |
| **Knowledge on Iron Folic Acid** |  |  |
| Good Knowledge IFA | 183 | 60.4 |
| Poor Knowledge IFA | 120 | 39.6 |

**Table 4:**
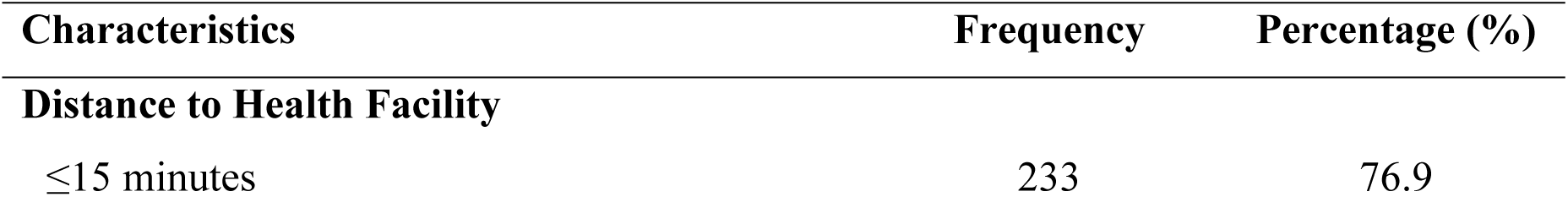
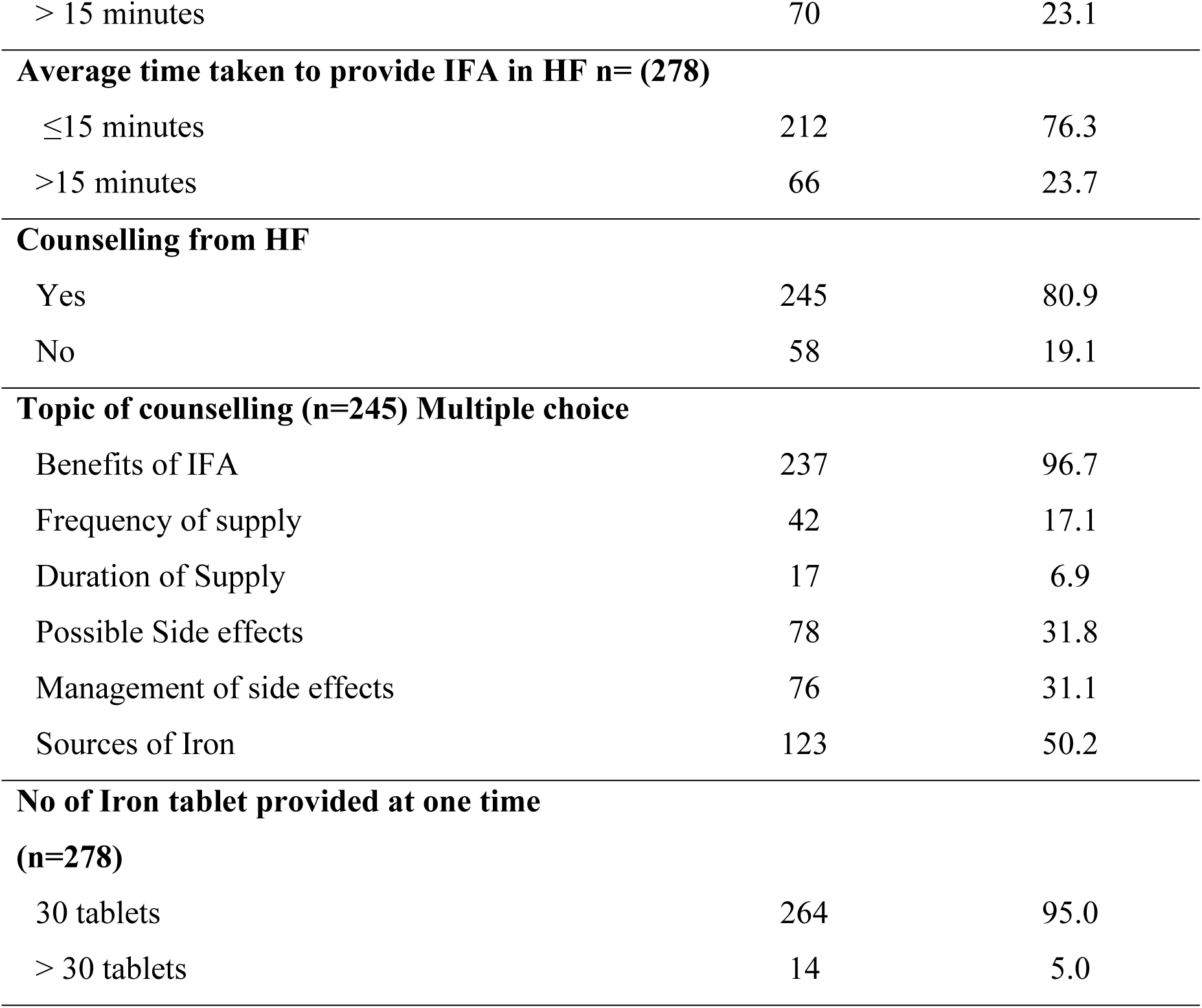
Health Facility Related Characteristics.

**Table 5:** Supplement related Characteristics.

| Characteristics | Frequency | Percentage (%) |
| --- | --- | --- |
| **Consumption of Iron Folic Acid** |  |  |
| Yes | 278 | 91.7 |
| No | 25 | 8.3 |
| **Started Trimester for consuming IFA (n=278)** |  |  |
| Second Trimester | 277 | 99.6 |
| Third Trimester | 1 | .4 |
| **Time of day (n=278)** |  |  |
| Morning | 1 | 0.4 |
| Evening | 54 | 19.4 |
| Night | 223 | 80.2 |
| **Side effects (n=278)** |  |  |
| Yes | 88 | 31.7 |
| No | 190 | 68.3 |
| **Types of Side effect\*\*(n=88)** |  |  |
| Constipation | 27 | 30.6 |
| Stool Discoloration | 3 | 3.4 |
| Vomiting | 79 | 89.7 |
| Abdominal Cramps | 9 | 10.2 |
| **Management of Side effects (n=88)** |  |  |
| Yes | 29 | 33 |
| No | 59 | 67 |
| **Ways of Management (n=29)** |  |  |
| Report to health care provider | 29 | 100.0 |

**\*\* Multiple choice**

### Health Facility Related

Majority of participants resided within a convenient distance to health facilities, with (76.9%) living less than or equal to 15 minutes away. Most respondents found the average time taken to receive iron folic acid (IFA) supplements at health facilities efficient, with (76.3%) receiving them within 15 minutes. Regarding counseling services provided by health facilities, majority of the respondents reported receiving it (80.9%), primarily focusing on the benefits of IFA (96.7%), information about possible side effects (31.8%), and their management (31.1%). Furthermore, the majority of participants received 30 tablets of iron at one time (95.0%), ensuring a consistent supply.

### Supplement Related Characteristics

The study findings reveal a significant uptake of iron folic acid (IFA) supplements among pregnant women, with over nine out of ten participants reporting consumption (91.7%), mainly starting during the second trimester (99.6%) when fetal development peaks. However, nearly one in three respondents experience side effects (31.7%), like vomiting (89.7%), constipation (30.6%) with only one in three seeking help (33%).

Table 6 shows the compliance of Iron Folic Acid Supplementation among the postpartum mothers of Bharatpur Metropolitan. Overall, compliance with Iron Folic Acid (IFA) supplementation among the respondents was determined to be 48.2%. Compliance rates were notably higher during the pre-pregnancy period at 75.2%, compared to the postpartum period at 48.2%. Moving to the reasons behind consistent IFA intake, the primary factor cited was the known benefits of iron folic acid (99.3%). Conversely, side effects (37.1%) and forgetfulness (34.8%) emerged as the primary reasons for irregular intake. A small proportion of respondents (8%) reported not consuming IFA throughout pregnancy, attributing this to a perceived lack of necessity.

**Table 6:** Compliance to Iron Folic Acid Supplementation.

| Characteristics | Frequency | Percentage (%) |
| --- | --- | --- |
| **Compliance to Iron Folic Acid** |  |  |
| Yes | 146 | 48.2 |
| No | 157 | 51.8 |
| **Compliance to IFA in pregnancy** |  |  |
| Yes | 228 | 75.2 |
| No | 75 | 24.8 |
| **Compliance to IFA after delivery** |  |  |
| Yes | 146 | 48.2 |
| No | 157 | 51.8 |
| **Reasons for regular intake of IFA\*\* (n=146)** |  |  |
| Known Benefits of Iron Folic Acid | 145 | 99.3 |
| Received proper counselling from health facility | 26 | 17.8 |
| **Reasons for irregular intake of IFA\*\* (n=132)** |  |  |
| Side effects | 49 | 37.1 |
| Forgetfulness | 46 | 34.8 |
| Not felt important after delivery | 26 | 19.6 |
| Unaware about the postpartum importance of IFA | 18 | 13.6 |
| **Reason for not consuming Iron Folic Acid\*\* (n=25)** |  |  |
| Not felt necessary | 12 | 48.0 |
| Unaware about the importance of Iron Folic Acid | 14 | 56.0 |

**\*\* Multiple choice**

### Logistic Regression Analysis of the factors Associated with compliance to IFAS

Table 7 presents the outcomes of logistic regression analysis, highlighting variables with significant links to compliance with iron folic acid supplementation. Variables that demonstrated significance in Pearson’s chi-square test (p-value < 0.05) were included in the logistic regression analysis. The multicollinearity test was performed to assess correlations among independent variables. This was achieved using the variance inflation factor (VIF), which did not reveal multicollinearity issues. The unadjusted odds ratio and adjusted odds ratio were calculated for each independent variable that showed significant association in the chi-square test, utilizing the enter method.

**Table 7:** Binary Logistic Regression Analysis of Factors Associated with Compliance to IFAS.

| **Variables** | **Unadjusted OR**<br>**(95% CI)** | **p-value** | **Adjusted OR**<br>**(95% CI)** | **p-value** |
| --- | --- | --- | --- | --- |
| **Ethnicity** |  |  |  |  |
| Brahmin/Chettri | 3.180(1.987-5.087) | <0.001 | 0.926(0.408-2.10) | 0.853 |
| Others | 1 |  | 1 |  |
| **Religion** |  |  |  |  |
| Hinduism | 3.333(1.729-6.422) | <0.001 | 5.367(1.1731-16.636) | **0.004** |
| Non-Hinduism | 1 |  | 1 |  |
| **Education** |  |  |  |  |
| Cannot read and<br>write/Non formal<br>education | 1 |  | 1 |  |
| Literate | 17.443(6.758-45.024) | <0.001 | 2.130(0.30015.143) | 0.450 |
| **Occupation** |  |  |  |  |
| House wife/Agriculture | 1 |  | 1 |  |
| Others | 4.731(2.548-8.785) | <0.001 | 1.100(0.415-2.914) | 0.848 |
| **Husband Education** |  |  |  |  |
| Cannot read and<br>write/Non formal<br>education | 1 |  | 1 |  |
| Literate | 11.891(4.925-28.712) | <0.001 | 1.783(0.339-9.384) | 0.495 |
| **Husband Occupation** |  |  |  |  |
| Employed/Business | 3.884(2.411-6.258) | <0.001 | 0.971(0.442-2.134) | 0.942 |
| Others | 1 |  | 1 |  |
| **Monthly Income** |  |  |  |  |
| NRs. ≤30000 | 1 |  | 1 |  |
| NRs.>30,000 | 7.215(4.315-12.063) | <0.001 | 1.924(0.848-4.366) | 0.942 |
| **Pregnancy Times** |  |  |  |  |
| Once | 2.128(1.344-3.370) | 0.001 | 0.907(0.418-1.971) | 0.806 |
| More than 1 time | 1 |  | 1 |  |
| **First ANC Months** |  |  |  |  |
| ≤ 3 months | 5.286(2.126-13.148) | <0.001 | 0.256(0.254-1.223) | 0.088 |
| > 3 months | 1 |  | 1 |  |
| **ANC Times** |  |  |  |  |
| <4 times | 1 |  | 1 |  |
| ≥4 times | 9.670(5.111-18.295) | <0.001 | 3.465(1.366-8.792) | **0.009\*** |
| **Health Facility for ANC** |  |  |  |  |
| Health<br>post/Governmental<br>Hospital | 1 |  | 1 |  |
| Private Hospital/Clinic | 12.281(6.539-23.441) | <0.001 | 2.194(0.857-5.615) | 0.101 |
| **Blood test during ANC** |  |  |  |  |
| Yes | 37.120(5.000-275.56) | <0.001 | 1.910(0.106-34.250) | 0.661 |
| No | 1 |  | 1 |  |
| **Miscarriage** |  |  |  |  |
| Yes | 4.597(1.270-16.636) | 0.02 | 17.984(0.719-450.08) | 0.79 |
| No | 1 |  | 1 |  |
| **Complication in pregnancy** |  |  |  |  |
| Yes | 2.675(1.405-5.093) | 0.003 | 2.274(0.869-5.955) | 0.94 |
| No | 1 |  | 1 |  |
| **Planned Pregnancy** |  |  |  |  |
| Yes | 34.252(4.605-254.75) | 0.001 | 4.590(0.326-64.587) | 0.259 |
| No | 1 |  | 1 |  |
| **Knowledge on Anemia** |  |  |  |  |
| Good Knowledge | 14.815(8.317-26.390) | <0.001 | 5.554(2.485-12.415) | **<0.001** |
| Poor Knowledge | 1 |  | 1 |  |
| **Knowledge on Iron Folic Acid** |  |  |  |  |
| Good Knowledge | 13.188(7.293-23.848) | <0.001 | 2.442(1.064-5.605) | **0.035** |
| Poor Knowledge | 1 |  | 1 |  |
| **Distance to Health Facility** |  |  |  |  |
| ≤ 15 minutes | 5.228(2.755-9.918) | <0.001 | 2.034(0.725-5.709) | 0.17 |
| >15 minutes | 1 |  | 1 |  |
| **Average time to collect IFA in HF** |  |  |  |  |
| ≤ 15 minutes | 3.079(1.714-5.531) | <0.001 | 1.484(0.600-3.671) | 0.393 |
| > 15 minutes | 1 |  | 1 |  |
| **Counselling** |  |  |  |  |
| Yes | 39.921(9.523-167.35) | <0.001 | 1.310(0.197-8.713) | 0.484 |
| No | 1 |  | 1 |  |
| **Total no of IFA tablets provided at one visit** |  |  |  |  |
| 30 tablets | 1 |  | 1 |  |
| >30 tablets | 5.821(1.278-26.515) | 0.023 | 2.040(0.331-12.55) | 0.442 |

*\*Statistically significant*

The variables that were significant in bivariate analysis were: ethnicity, religion, education, occupation, husband education, husband occupation, monthly income, pregnancy time, times of ANC Visit, First ANC month, place of ANC, blood test in pregnancy, history of anemia, planned pregnancy, history of abortion, history of miscarriage, complication in pregnancy, distance to health facility, average time to collect IFA tablets, counselling, knowledge on anemia, knowledge on iron folic acid, total number of IFA tablets provided at one visit.

The adjusted odds ratio for individuals of the Hindu religion was 3.333 (CI: 1.729-6.422), suggesting that individuals in this group had over three times higher odds of complying with Iron Folic Acid (IFA) supplementation. Likewise, the adjusted odds ratio for participants attending more than four ANC visits was 3.465 (CI: 1.366-8.792), indicating that participants with more than four ANC visits were over three times more likely to comply with IFA supplementation. Similarly, participants with good knowledge of IFA supplementation had five times higher odds of complying compared to those with poor knowledge. Moreover, the adjusted odds ratio for participants with good knowledge of IFA supplementation was 13.188 (CI: 7.293-23.848), indicating that participants with good IFA knowledge were over 13 times more likely to comply with supplementation.

## Discussion

### Compliance with Iron Folic Acid Supplementation

This study aims to assess the compliance of iron folic acid supplementation and its associated factors among postpartum mothers of Bharatpur Metropolitan City, Chitwan, Nepal. The findings of our research revealed that almost half of the respondents (48.2%) adhered to Iron Folic Acid (IFA) Supplementation. These results are consistent with a previous study in Wondo District, which showed a 44.3% compliance rate, and another study in Debre Tabor General Hospital, Ethiopia, which reported a 44% compliance rate [4,10] This moderate level of compliance indicates that these areas might share similar challenges and facilitators for promoting IFA supplementation adherence.

In contrast, a study in Kathmandu found a significantly higher compliance rate of 78.6% [11]. This difference could be attributed to various factors, such as better healthcare infrastructure and more comprehensive antenatal care programs in urban settings like Kathmandu. Additionally, the Nepalese government’s effective communication and enforcement of IFA tablet guidelines in urban areas might lead to higher adherence rates.

In the Metema district, the compliance rate was notably lower at 34% [12].This lower rate could be due to limited healthcare access, lower quality antenatal care, and less effective health education programs. Socio-economic challenges and cultural practices might also contribute to lower adherence.

The high compliance rate of 81.74% in West Bengal highlights the success of effective healthcare delivery and education in the region [13]. The use of multistage sampling in the study might have provided a comprehensive view of adherence behaviors across different sub-centers, showcasing strong health infrastructure and targeted interventions. Compared to our study, West Bengal’s strategies in health education and service delivery could serve as a model for improving adherence in other regions.

Additionally, our study found that the primary reason for non-compliance with IFA Supplementation was a lack of awareness about its postpartum importance. Other major factors included forgetfulness and experiencing side effects. Conversely, the known benefits of IFA Supplementation for both mother and child were the main predictor of compliance among respondents.

### Factors associated with compliance to Iron Folic Acid Supplementation

The frequency of Antenatal Care (ANC) visits has emerged as a crucial factor influencing adherence to Iron Folic Acid (IFA) supplementation, indicating that participants attending more than four ANC visits are more likely to adhere to IFA supplementation guidelines. This finding aligns with several other studies, which have similarly emphasized the positive association between the number of ANC visits and IFA compliance. For instance, research in Ethiopia has shown that mothers who attended four or more three times more likely to comply with IFA supplementation compared to those with fewer visits [14]. Consistent results were also observed in studies conducted in Indonesia [15], Sub-Saharan African countries [16], Mulago National Referral Hospital in Uganda [17], and Southern Senegal, Amhara [18]. However, a study among pregnant women at an antenatal care clinic at Sub District Hospital in Ballabgarh [19] and study conducted in Wondo District, Omoria, Ethiopia found no significant association between ANC visit frequency and IFA compliance [4].

The consistency of positive associations in various settings can be attributed to several factors. Frequent ANC visits offer multiple opportunities for healthcare providers to educate and motivate pregnant women about IFA supplementation, enhancing adherence. Additionally, women attending more ANC visits typically receive consistent and comprehensive care, including ongoing monitoring and support, which reinforces adherence to supplementation guidelines. However, the contrasting finding may be due to contextual differences.

Compliance with Iron Folic Acid (IFA) supplementation was found to be higher among women with good knowledge of anemia. Our study suggested that knowledgeable participants were more than five times as likely to comply with IFA supplementation, underscoring that a strong understanding of anemia is a robust predictor of compliance, even when considering other variables. This finding aligns with studies conducted in Kenya and the Goba district of Ethiopia [4,20].Furthermore, another cross sectional study conducted on postnatal mother in Kenya also identified the positive association between the knowledge on anemia and compliance to IFA [21]. This likely reflects that pregnant women with satisfactory knowledge are more aware of the adverse consequences of iron and folic acid deficiencies on maternal and fetal health, thereby increasing their efforts to adhere to supplementation guidelines.

Conversely, a cross-sectional study in Soro district, Hadiya Zone, Ethiopia, did not show any association between knowledge and adherence to IFA supplementation [22]. Furthermore, another study conducted among pregnant and postnatal women in a hospital in Kathmandu, Nepal found no association between the level of knowledge and compliance with iron and folic acid consumption[23]. These discrepancies could be attributed to several factors. In some contexts, structural and systemic barriers such as lack of access to supplements, economic constraints, or poor quality of healthcare services may impede adherence despite good knowledge.

In our study Participants with good knowledge of Iron Folic Acid (IFA) supplementation were more likely to comply with IFA supplementation compared to those with poor knowledge, indicating that participants with good IFA knowledge were over 13 times more likely to comply with supplementation. This result is highly statistically significant, reflecting a strong direct association between IFA knowledge and compliance. The association between knowledge of IFA and compliance has been examined in several studies. In a cross-sectional study conducted on pregnant mothers attending governmental health institutions in Adwa town, Tigray, Ethiopia, knowledge about IFA supplementation was identified as a factor associated with adherence to the supplement [24]Pregnant women with satisfactory knowledge of IFA supplementation were found to have twice the odds of adherence compared to those with unsatisfactory knowledge. This finding aligns with studies conducted in Kenya and Goba district, Ethiopia [25,26].

In summary, existing evidence indicates a positive association between knowledge about IFA supplementation and adherence to the regimen. This suggests that being well-informed about IFA could play a crucial role in influencing compliance with IFA supplementation.

However, a cross-sectional study in Soro district, Hadiya Zone, Ethiopia, did not show any significant relationship between knowledge of IFA supplementation and compliance [22]. Similarly, a study conducted in Dangila, Northern Ethiopia, also showed no relationship between education on IFA tablets and compliance with IFA supplementation [27]. These contrasting findings may be due to differences in study populations, healthcare infrastructure, and the effectiveness of educational interventions. Understanding these variations is important for designing effective strategies to improve IFA supplementation adherence.

### Strength and limitation

This study represents the first community-based study conducted in the study area, focusing on adherence to iron folic acid supplementation and identifying the factors influencing this compliance. Additionally, to ensure proper compliance, IFA tablets should be consumed for 45 days after delivery. Therefore, our study included women who completed their 45days postpartum period.

One significant limitation of our study is the use of self-reports to evaluate noncompliance with oral IFA supplementation, which may result in either underestimation or overestimation by participants, thereby introducing reporting bias. Furthermore, the potential for recall bias and social desirability bias exists when participants report their compliance. This research focuses on the users’ perspective regarding adherence to iron folic acid supplementation. However, it does not identify the factors contributing to noncompliance from the health service providers’ perspective.

## Conclusion

The study highlights a significant gap in the compliance rate of Iron Folic Acid Supplementation (IFAS) among postpartum mothers in Bharatpur Metropolitan City, with only about half of the participants adhering to the supplementation guidelines. Despite various government initiatives aimed at improving IFAS adherence, the compliance remains low, underscoring the need for more effective interventions.

The study identifies key factors influencing IFAS compliance, including religion, frequency of ANC visits, and knowledge about anemia and IFAS. The primary reason for non-compliance is a lack of awareness regarding the importance of IFAS during the postpartum period, which points to a critical area for intervention.

To address the identified barriers and improve compliance with IFAS, the study suggests several strategies. These include creating educational programs that dispel myths and emphasize the benefits of IFA supplements during pregnancy and the postpartum period, and engaging religious leaders to ensure cultural sensitivity. Additionally, mobilizing community health workers for home visits can help reinforce the importance of IFAS and monitor compliance. Future research should also incorporate healthcare providers’ perspectives to gain a more comprehensive understanding of IFAS compliance dynamics. Implementing these recommendations can lead to a more effective approach in promoting IFAS adherence, ultimately contributing to better health outcomes for mothers and children in the community.

## Data Availability

All relevant data are within the manuscript and its Supporting Information files.

## Acknowledgement

The authors extend their gratitude to the School of Health and Allied Sciences, Faculty of Health Sciences, Pokhara University. Additionally, heartfelt thanks are extended to the health and Promotion Section as well as the health facilities of Bharatpur Metropolitan City for granting permission to carry out the study. Lastly, we would like to convey our profound gratitude to the study participants who generously contributed their time and provided valuable information for this research.

## Author Contribution

Conceptualization: Amshu Pokhrel (AP), Bimala Bhatta (BB)

Data Curation: Amshu Pokhrel (AP), Anup Adhikari (AA)

Formal Analysis: Amshu Pokhrel (AP), Bimala Bhatta (BB), Anup Adhikari (AA)

Investigation: Amshu Pokhrel (AP)

Methodology: Amshu Pokhrel (AP), Bimala Bhatta (BB)

Writing – Original draft: Amshu Pokhrel (AP)

Writing – review and editing: Bimala Bhatta (BB), Anup Adhikari (AA)

